# How Americans encounter guns: Mixed methods content analysis of YouTube and internet search data

**DOI:** 10.1101/2022.07.15.22276403

**Authors:** Megan L. Ranney, Frederica R. Conrey, Leah Perkinson, Stefanie Friedhoff, Rory Smith, Claire Wardle

## Abstract

Firearm-related injury and death is a serious public health issue in the U.S. As more Americans consume news and media online, there is growing interest in using these channels to prevent firearm-related harms. Understanding the firearm-related narratives to which consumers are exposed is foundational to this work.

This research used the browsing behavior of a representative sample of American adults to identify seven naturally occurring firearm-related content ecosystems on YouTube, and the demographics and internet search patterns of users affiliated with each ecosystem.

Over the 9-month study period, 72,205 panelists had 16,803,075 person-video encounters with 7,274,093 videos. Among these, 282,419 videos were related to firearms. Using fast greedy clustering, we partitioned users and channel interactions into seven distinct channel-based content ecosystems that reached more than 1/1000 YouTube users per day. These ecosystems were diverse in reach, users, and content (e.g., guns for self-protection vs. guns for fun).

On average, 0.5% of panelists performed a firearm-related internet search on a given day. The vast majority of these searches were related to mass shootings or police-involved shootings (e.g., “active shooter”), and virtually none were about more common firearm harm such as suicide. Searches for firearm safety information were most common among panelists affiliated with the “Hunting & Fishing” and “Guns & Gear” ecosystems, which were watched primarily by older, white men.

These findings identify an opportunity for analyzing firearm-related narratives and tailoring firearm safety messaging for users affiliated with specific online content ecosystems.

**Highlights:**

- We used a mixed methods network analysis of YouTube users’ viewing patterns and identified seven distinct firearm-related content ecosystems.
- Ecosystems vary by reach, audience demographics, and audience firearm-related search patterns.
- The ecosystems contain a wide variety of narratives about firearm use, safety, and potential harms.
- Understanding the diverse narratives across these ecosystems, their respective audiences and audience search patterns can inform future work to reduce firearm-related harms and increase safety.

## 1. Introduction

Firearm-related injury and death are a major public health problem, with more than 40,000 deaths and an estimated 85,000-130,000 injuries a year (CDC, 2021). Firearm injury is now the leading cause of death for American youth age 1-19, the most common mechanism of death from intimate partner violence, and the most common mechanism of suicide death (Andrews et al., 2022; CDC, 2021). Young Black men are disproportionately more likely to be victims of firearm homicide, while older White men are at highest risk of firearm suicide (Kaufman & Richmond, 2020; Riddell et al., 2018). Rising rates of firearm injury and death since 2020 have been accompanied by a reported increase in the proportion of Americans who own a firearm (Gun Violence Archive, n.d.-b; Kaufman et al., 2021). Prior to COVID-19, 40% of Americans were firearm owners, and preliminary data suggests that another 2.9% became first-time owners during the first year of the pandemic (Anestis et al., 2021; Miller et al., 2022).

Shifting norms and behaviors regarding firearm “safety” and “harm” may be among the reasons for these increases in American firearm ownership, injury, and death. Subjective beliefs about the risk of firearm harm, and perceived ability to control that harm, are informed by factors ranging from in-person social networks to the media. Indeed, extensive evidence supports that one’s likelihood of firearm injury depends on patterns in one’s in-person social network (Papachristos & Wildeman, 2014). Qualitative studies of small groups of firearm owners suggest that so-called “gun culture” influences firearm ownership choices, firearm safety behaviors and firearm injury risk (Thomas et al., 2022; Yamane, 2017; Yamane et al., 2020, 2021) - although the determinants of this culture remain poorly defined. Unfortunately, data from firearm retailers suggest that the majority of new firearm owners did not receive basic safety training, despite ample evidence that consistent safe storage and safe handling are key to prevention (NSSF, 2022; Rowhani-Rahbar et al., 2016). Other preliminary work suggests that misleading and selective reporting by the media about firearm injury may lead to the American public making inaccurate judgements about the most common firearm injury risks (Jetter & Walker, 2018; Kaufman et al., 2020) and the availability of prevention strategies (Pallin et al., 2021).

As more Americans rely on online and social networking sites for news and information, it is increasingly likely that these spaces - and the words and narratives included in them - are an important driver of Americans’ current understanding of firearm injury, safety, and injury prevention (Betz et al., 2021; Walker & Matsha, 2021). For example, Twitter conversations in Latin America have been demonstrated to over-represent the likelihood of violent crime, and to represent societal fears more than actual prevalence (Prieto Curiel et al., 2020). Specific types of content of Twitter conversations in Detroit and Chicago have been shown to drive community conceptions and patterns of firearm injury (Patton et al., 2017; Stuart et al., 2020). Similarly, qualitative analysis of comments about online firearm-injury-prevention news articles have been used to identify areas of agreement and tension between health professionals and firearm owners (Knoepke et al., 2017). To our knowledge, no one has used large online media datasets to empirically examine the spaces and ways in which a diverse sample of the American public encounters narratives about guns.

As a first step to understanding the nature of firearm-related content to which consumers are exposed in social media, we aimed to define the nature, prevalence, and risk-related firearm content on YouTube. We chose YouTube as it is the most commonly consumed online social media platform across demographic groups in the United States (Auxier & Anderson, 2021), and as proprietary, anonymized databases of YouTube watching data are available.

Specifically, the goals of this analysis were to, first, characterize distinct online firearm-related narrative typologies on YouTube; second, describe the demographic characteristics of subpopulations that consume content within each typology on YouTube; and third, understand how these distinct audiences interact with firearm injury-related narratives in internet searches.

## 2. Methods

### 2.1 Data sources

#### 2.1.a: YouTube

We obtained data about YouTube consumption from December 1, 2020-August 31, 2021 from a proprietary, representative research panel of American adults’ desktop and laptop browsing behavior maintained by a major media measurement firm. Because panelists can opt in and out of the panel, we use person-days as a reliable means of measuring total panel volume. The measurement panel maintains records of self-reported user demographics (age, race, and gender) from which we calculated overall demographics for all people watching any YouTube content and demographics for people watching any firearms-related YouTube content.

“Videos” were defined as individual videos on the platform. “Channels” were identified as per standard YouTube practice.

We retrieved video details including channel title, title, description, and creator-assigned “tags” or descriptors for all the videos in this sample from the YouTube API. Not all video details were available at the time of retrieval. We determined whether videos were considered relevant to “guns” or “firearms” if any content in the title, description, or tags matched this regular expression (note that ‘\y’ represents a word boundary to the Postgres database we used):

*‘\ygun\y*|*\yguns\y*|*\yfirearm\y*|*\yfirearms\y*|*\yshooting\y*|*\yammo\y*|*\yammunition\y*|*\yrifle\y*|*\ypisto l\y*|*\yshotgun\y*|*\yrevolver\y*|*\yautocannon\y*|*\yactive shooter\y’*

AND if none of the text matched this regular expression (to exclude videos that are popular culture icons but not directly related to firearms and gun culture):

*‘top gun*|*god family and guns*|*roses*|*kelly*|*glue*|*staple*|*nail*|*heat gun*|*gloves up*|*massage*|*shooting star*|*outdoor shooting*|*shooting for*|*basketball shooting*|*shooting video*|*shooting to*|*trouble shooting*|*shooting outside*|*\yarrow\y*|*shooting guard*|*tape gun*|*smoke gun*|*smoking gun*|*impact gun*|*bullet train*|*black rifle coffee*|*bullet journal*|*shotgun red*|*thera*|*photo*|*\ycum\y*|*shooting up*|*bullet point*|*shooting pain*|*video shooting*|*caulk*|*paint gun*|*shotgun mic*|*signal gun*|*clothespin gun’*

#### 2.1.b Internet searches

As a preliminary examination of types of firearm harm information related to YouTube ecosystems, we retrieved all internet search queries submitted to google.com, duckduckgo.com, and yahoo.com by panel members during the study period, using the search terms above. This method eliminates seasonality, weekly cycles, and big spikes associated with one time events.

Searches relevant to “firearm safety” matched the regular expression query: ‘safe|training|\\yclass\\y|permit|license|prevent’. Searches relevant to “firearm policy” matched the query: ‘law|control|bill|legal’. To identify searches relevant to other firearm topics, we coded 250 unsupervised clusters of these search terms for relevance to firearm harm or injury. Specifically, we embedded the terms using a pretrained [sentence transformer](https://huggingface.co/sentence-transformers/msmarco-MiniLM-L12-cos-v5). Using an a priori objective of 400 searches per cluster, we clustered the resulting vectors into 250 clusters using R’s [kmeans](https://www.rdocumentation.org/packages/stats/versions/3.6.2/topics/kmeans) procedure.

### 2.2 Analyses

#### 2.2.1 YouTube social network, qualitative, and demographic analysis

To define the specific networks of YouTube narratives, we considered all firearm-relevant YouTube video encounters identified through the YouTube search. We treated each distinct YouTube channel (not individual video) as a node in a network, and created edges between channels that were watched by the same person on the same day. For example, if a single user watched a firearm-related video on CNN and a second video on Fox News on the same day, we created an edge between the CNN and Fox News nodes. Each pair was represented in the model a maximum of one time per day (to represent channel reach rather than viewing frequency). Importantly, we do not know whether channel choice was user-driven or driven by the YouTube recommendation algorithm; we only know that these channels were watched by the same individuals on the same days.

We then partitioned the network (with edge weights representing the number of distinct person-days on which an edge appeared) into “ecosystems” using the fast greedy clustering from the [igraph](https://igraph.org/) package in R (Clauset et al., 2004; v1.2.8; Csardi & Nepusz, 2006). *A priori*, we decided to retain all ecosystems with average daily reach of firearm-related content among YouTube users of 1 out of 1,000 users or greater.

We mapped individual channels’ importance in each retained ecosystem using both reach (proportion of users reached per day) and proximity of channel nodes. To characterize the content of each ecosystem, 2 researchers each watched up to 3 videos by reach (only videos that reached more than one user during the study period were included) in each of the top 15 channels of each ecosystem (by closeness) and independently characterized the content. In discussion, we refined the ecosystem characterizations, and identified potential exceptions to the primary ecosystem themes.

We also calculated descriptive statistics, weighted to represent the US population based on 2020 decennial Census and American Community Survey estimates, to examine the characteristics of each ecosystem’s users, using panelists’ self-reported demographics as compared to average YouTube users’ reported demographics (U.S. Census Bureau, 2021, 2022). We did not test for statistical significance given the large quantity of data, in congruence with our prior work with these datasets (Allen et al., 2020).

#### 2.2.2 Internet search cluster analysis

From each of the internet search clusters we extracted six randomly selected examples. These were evenly split between examples assigned to the cluster that were close to the centroid, and examples assigned to the cluster that were far from the centroid or relatively poorly described by the cluster. Two researchers manually examined both “good” (close to centroid) and “bad” (far from centroid) examples to confirm whether clusters were relevant (“good”), not relevant (“bad’), or ambiguous. The two researchers manually coded each cluster’s examples and top keywords for relevance to human firearm injuries (including suicide, accidents, homicides, or mass shootings). All clusters were double-coded, and differences were resolved through discussion amongst the team.

For each day, we calculated the percent of people in the panel who searched anything firearm related, and then averaged these percentages across all days. As a sensitivity analysis, we conducted these analyses both with and without “ambiguous” clusters.

### 2.3 Ethical Compliance

HarmonyLabs served as the data curator and analyzer of data. Consistent with their prior work, Internet search and YouTube consumption data is completely anonymous and has been stripped of all potential PII (Personally Identifiable Information). YouTube metadata which we joined to the panel data is publicly available and contains no PII or sensitive information. Panelists have opted into tracking, have been compensated for their contributions, and may opt out at any time. The team conformed with all privacy, confidentiality, and ethical requirements of the panel’s media measurement company.

## 3. Results

Between December 1, 2020 and August 31, 2021, the opt-in panel included 2,111,309 person-days (measuring behavior of a total of 72,205 distinct American adults) of identifiable videos consumed on YouTube. The analysis considered descriptions for 16,803,075 person-video encounters with 7,274,093 distinct videos. Of these, 282,419 videos were identified as related to firearms using our search terms. On an average day, 7% of people in the panel who watched any YouTube content consumed some content relevant to firearms.

Partitioning the network of user and channel interactions yielded 7 ecosystems that had a reach greater than 1 per 1000 YouTube users per day (see **Table 1** & **Figure 1**). An ecosystem might contain channels that have high-reaching content unrelated to guns (i.e., Movies or Music), but a lower reach of gun-specific content. These ecosystems were qualitatively described as:

1. *Hunting & Fishing* (characterized by celebration of firearms and sport, treating firearms as both a hobby and a pedagogy). The stories in this ecosystem tend to feature air rifles specifically and are about shooting for sport or shooting animals, but not shooting people.
2. *Gaming* (online or video-gaming). a highly social and exclusively virtual ecosystem. While it includes discussions of technical specifications of firearms, this gaming community places more emphasis on firearms being used to connect with others through shared enjoyment.
3. *Movies* (representative of mainstream imagination and big-screen narratives). Much of the content in this ecosystem comes from Movieclips; it is all creative and imaginative imagery.
4. *Guns & Gear* (featuring guns themselves as the subjects). All of the videos in this ecosystem are explicitly about firearms, and the ecosystem’s language is characterized by notions of “self-defense,” safety, and responsibility. The ecosystem’s videos’ goals are explicitly about shooting (lethally or non-lethally) other people.
5. *Guns 4 Fun* (recreational and simulation real-life “battle” games, such as Nerf gun wars). This ecosystem mostly relates to mock guns, including military simulation airsoft games and Nerf games simulating Fortnite gaming battles. The tone of this ecosystem is playful and fun. These creators celebrate their weaponry but clearly know the weapons are pretend.
6. *Music* (primarily metaphorical use of gun imagery and words). While many of the gun references in popular music are highly metaphorical or referential, much of the top content in this ecosystem centers real gun violence either from the perspective of the victim of violence.
7. *News & Hot Takes* (discussions of current events). The tone of gun-related content in News & Hot Takes is one of danger paired with combative arguments between partisan extremists. Videos move from recent reports of national shootings on local news to angry political rants for and against gun control. Language about safety, responsibility, or enjoyment is largely missing.

**Table 1:** Qualitative and quantitative descriptors of the seven naturally occurring firearm narrative ecosystems on YouTube. “Reach” is defined as the average number of YouTube users who interact with channels in the ecosystem each day.

| **Ecosystem** | **Description** | **Examples of Channels** | **Reach** |
| --- | --- | --- | --- |
| Hunting & Fishing | Videos characterized by their celebration of firearms as sport, featuring stories about shooting as a hobby. Videos often describe shooting animals, but not people. | americanairgunhunter; Fieldsports Channel; Joel Goodz; South Florida Fishing; Trotliners | ~1/1,000 |
| Gaming | Highly social and exclusively virtual, videos in this ecosystem place an emphasis on entertainment, collaboration, and connecting with others via gaming communities and shared enjoyment. | Aztecross; GameGrumps; Gus Johnson; LetsPlay; teamfortress | ~5/1,000 |
| Movies | Videos in this ecosystem come from movies and adjacent content and are primarily creative in nature: inventive, dramatic, and only incidentally dangerous. | BeltFeds.Com LLC; BurdenWorld; Grjngo - Western Movies; Miami Vice; Movieclips | ~7/1,000 |
| Guns & Gear | Videos which include guns themselves as the subjects and characterized by notions of self-defense, safety, and responsibility. The explicit goal of most videos is preparing and training for shooting (lethally or non-lethally) other people. | AtlanticFirearms; Cheaper Than Dirt!; Forgotten Weapons; GunsAmerica; Jerry Miculek - Pro Shooter | ~9/1,000 |
| Guns 4 Fun | One of two distinct gaming ecosystems, videos in this category feature mock guns, military simulation airsoft games, and nerf games, with much of the content containing a playful and fun tone. | Aaron Esser; AleksWorkshop; NICKMERCs; Silo Entertainment | ~10/1,000 |
| Music | The highest reach entertainment ecosystem, many of the gun references in popular music are highly metaphorical or referential. Much of the content centers on real “gun violence” (shootings of other people), either from the perspective of the victim or the perpetrator. | ArianaGrandeVevo; Blake Shelton; EminemMusic; Lil Baby Official 4PF; Queen Official | ~12/1,000 |
| News & Hot Takes | The tone of gun-related content in this ecosystem is one of danger paired with combative arguments between partisan extremists, often consisting of reports of national shootings on local news, and angry political rants for and against gun control. | ABC News; CNN; Fox News; Guardian News; StevenCrowder | ~15/1,000 |

**Figure 1:**
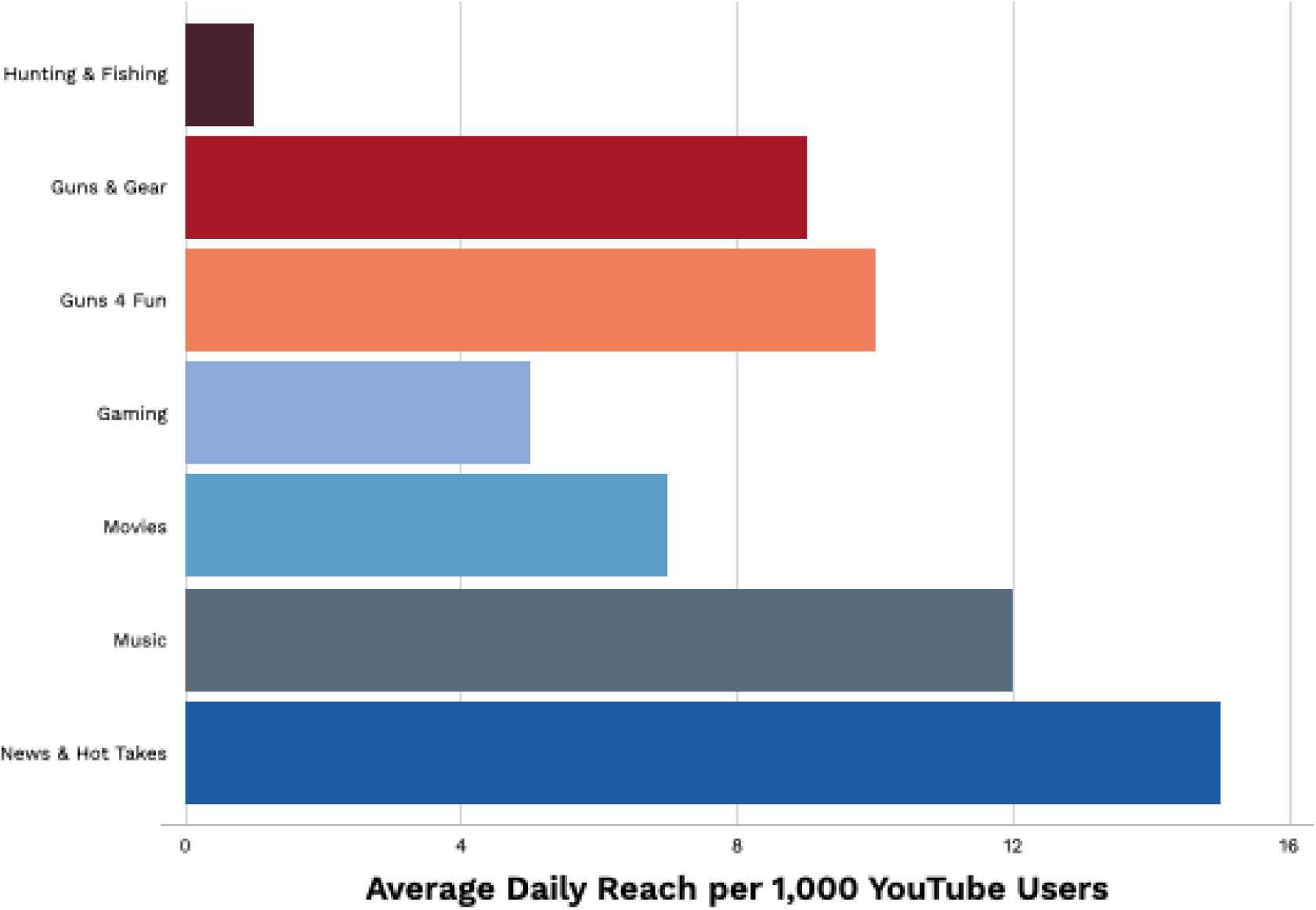
The average daily reach across the seven naturally occurring firearm narrative ecosystems on YouTube.

Demographic differences were observed between the users of the ecosystems. For example, with the exception of the Music ecosystem, consumers of all “gun” ecosystems were disproportionately male compared with the average YouTube user. The “Hunting and Fishing” and “Guns and Gear” ecosystems were more often White Non-Hispanic than the average YouTube user (70% and 81%, respectively); these ecosystems were also older than the average YouTube user. In comparison, the “Guns 4 Fun” and “Music” ecosystems were more often Black, and the “Guns 4 Fun” and “Gaming” ecosystems were more often age less than 30 (42% and 62%). (See **Table 2**.)

**Table 2:** Average demographics of YouTube users overall, and in specific firearm narrative ecosystems. % of YouTube users in a given ecosystem who fit a given demographic category (drawn from market panel data); exact number not available

|  | **All**<br>**YouTube** | **Hunting**<br>**&**<br>**Fishing** | **Guns &**<br>**Gear** | **Guns 4**<br>**Fun** | **Games** | **Movies** | **Music** | **News &**<br>**Hot**<br>**Takes** |
| --- | --- | --- | --- | --- | --- | --- | --- | --- |
| **Age:** |  |  |  |  |  |  |  |  |
| *Over 50* | 38% | 47% | 56% | 15% | 12% | 37% | 26% | 41% |
| *Under 30* | 27% | 20% | 16% | 42% | 62% | 33% | 33% | 24% |
| **Race/Ethnicity:** |  |  |  |  |  |  |  |  |
| *Non-Hispanic White* | 62% | 70% | 81% | 61% | 63% | 66% | 61% | 65% |
| *Black* | 14% | 11% | 7% | 19% | 10% | 13% | 18% | 16% |
| *Hispanic* | 16% | 9% | 6% | 14% | 17% | 13% | 16% | 12% |
| **Gender:** |  |  |  |  |  |  |  |  |
| *Male* | 51% | 75% | 84% | 58% | 76% | 72% | 50% | 69% |

Examining the internet searches used by all internet users and by people in the seven content ecosystems, we identified 103,727 internet searches conducted by panel members during this period.

Using the methods described above to identify firearm-related searches, after initial coding, nine of 250 potential firearm-related clusters were deemed irrelevant to firearm harm or injury (e.g., things like “[revolver news](https://www.revolver.news/)”). Forty-five were clearly and exclusively related to firearm harm and injury; 193 were about firearms, but unrelated to harms; and three were ambiguously related to firearm harms (including one that contained searches about both threatening people with guns and also using firearms in biathlon events). For instance, the six examples examined for one cluster were:

|  |  |
| --- | --- |
| Close to centroid | shooting in nova scotia |
| Close to centroid | nova scotia shooting |
| Close to centroid | see jacob blake shooting |
| Far from centroid | tampa bay shooting |
| Far from centroid | shooting cape girardeau mo |
| Far from centroid | ryan le-nguyen released on bond after-shooting-a bc over a tricycle dispute |

Since both the “good” and “bad” examples of this category were relevant to firearm harm and injury, we were confident that the entire cluster was relevant.

On average, 0.5% of panelists searched for something related to firearms on any given day. Of those, ∼4% searched for anything related to firearm policies (e.g., “gun control” or “laws”) and ∼3% searched for anything related to firearm safety (e.g., “safes” “training”). The most common categories of firearm related searches were information about firearm-related injuries or harms (e.g “active shooter”), of which 96% were about mass shooting events or police-involved shootings, and virtually none were about more common kinds of firearm harm such as suicide or unintentional injuries.

Different search terms mapped differentially onto the ecosystems. See **Figure 2** for specific words commonly contained in firearm-related searches associated with each ecosystem: for example, searches for “lyric” and “song” were contained in larger searches for specific songs about guns and violence, and “grain” was used in searches that refer to gunpowder specifications.

**Figure 2:**
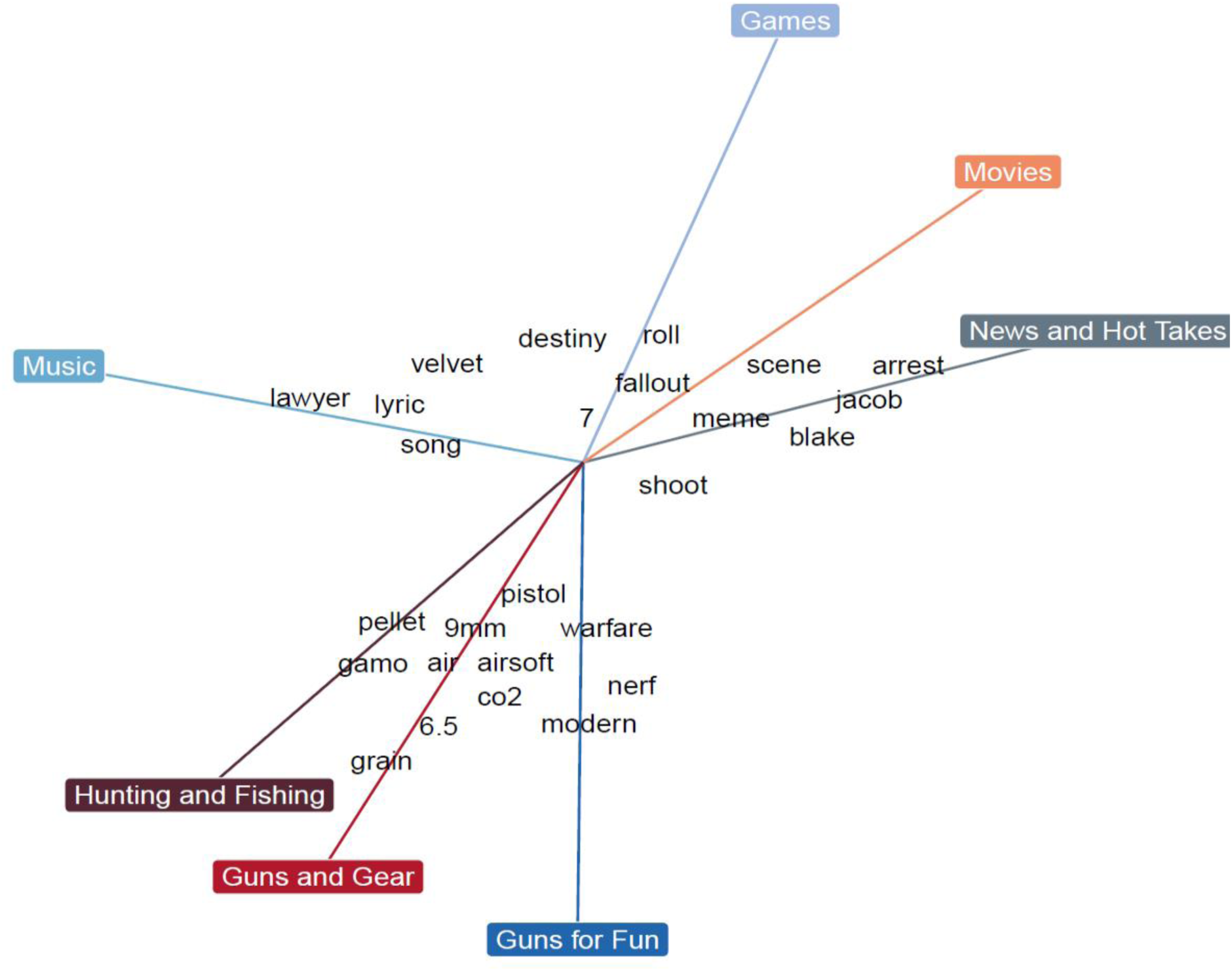
Spider diagram of common words contained in firearm-related searches associated with each firearm ecosystem. Words closer to ecosystem names were searched more often by same-day viewers of those ecosystems. These words are selected based on both frequency and distinctiveness with respect to the topics.

## 4. Discussion

By examining national YouTube video consumption data using a mixed-methods social network approach, we surfaced seven distinct naturally-occurring online ecosystems in which consumers are exposed to a variety of narratives about firearm use and safety. To our knowledge, this mixed-methods analysis is the first to provide data-driven insights into types of firearm-related narratives presented on YouTube, the demographic characteristics of groups who consume these narratives, and related internet search patterns. Our findings highlight the dynamic and pervasive nature of online social network discussions of and narratives about firearms. It also demonstrates gaps in, and the potential value of engaging the social media universe as part of efforts to improve on, Americans’ knowledge about and prevention of firearm harm.

Our study highlights that online social narratives - including YouTube - should be part of the discussion about who and how Americans learn about and conceive of firearms and firearm safety. According to national data, YouTube is used by 54% of its American adult users each day (Auxier & Anderson, 2021). In our analysis, nearly 7% of panelists encountered some firearm-related YouTube content each day. In other words, a substantial portion of American adults are likely exposed to online social media firearm narratives on a daily basis. The salience of narrative in influencing beliefs about, use of, and response to firearms and firearm injury have been amply demonstrated by others (McGinty et al., 2013; Yamane et al., 2020)(Gerbner & Gross, 1976; Iyengar, 1991). The importance of trusted messengers in conveying accurate information about firearm safety, harm, and injury prevention is also frequently discussed (Anestis et al., 2021; Betz et al., 2021). Our findings suggest that YouTube is a potentially important source of trusted messengers.

This study also provides a first glimpse into the wide diversity of narratives about firearms on YouTube. Quantitative and qualitative data on pre-COVID firearm ownership suggest there is more than one “gun culture” among firearm owners (Betz et al., 2021; Boine et al., 2020; Yamane, 2017). Our qualitative and quantitative examination of YouTube data and corresponding internet searches demonstrates that online cultural narratives around firearms, in general, are even more diverse than those suggested through studies of firearm owners - whether in reach, narrative content, and user demographics. Each of the seven narrative ecosystems had its own stories, users, and corresponding searches on firearm safety and harm.

For example, the small and self-contained “Hunting & Fishing” and “Guns & Gear” ecosystems - which were most focused on the actual use of firearms, largely for self-protection or as a hobby - were primarily watched by older, white men. Internet searches for firearm safety information were most commonly seen among panelists affiliated with these ecosystems; but, even here, discussions of the prevalence of, or risk factors for, the most common kinds of firearm harm were sparse. The demographics of these two ecosystems (and related searches) do not, of course, reflect the most quickly growing groups of gun owners - women and minorities (Miller et al., 2022; NSSF, 2022). In other words, although some firearm safety information is present, the spaces in which it shows up are limited and less diverse.

Indeed, despite popular attention to the issue of “misinformation” (Aspen Institute, 2021; Office of the Surgeon General, 2021), our analysis (while not focused specifically on identifying misinformation) suggests that the bigger issue in firearm injury prevention is *absence* of information in online narratives about firearms (Golebiewski & boyd, 2019). Throughout all seven ecosystems, we identified few if any spontaneous stories, much less corresponding internet searches, about either the ways in which guns harm people most commonly (i.e., suicide, unintentional injury, and partner violence) or how to maintain safety for one’s self and one’s family (outside of the self-protection narrative). Indeed, in the firearm narrative ecosystems frequented primarily by women, non-White, or younger YouTube users (i.e., “Games”, “Music,” “Guns 4 Fun”), searches and stories about safety were virtually non-existent. Future work should further describe the different kinds of firearm safety narratives and the sentiment and trust of those who provide those stories. Americans are simply not exposed to the personal, localized burden of, or risks for, firearm injury that dominate actual injury patterns. Our work therefore suggests that adding to existing narratives - rather than seeking to replace or out-shout them - may be a fruitful avenue for future work; it also reinforces the importance of tailoring to different ecosystems’ audiences.

For the majority of the YouTube ecosystems, firearms are incorporated into daily definitions of fun, social connection, and leisure, and not into politics. Although others’ work suggests that news and creative media have created a polarizing narrative about firearms (Bonnevie et al., 2020; Dada et al., 2022; Quinn & Andrasik, 2021), in our analysis, polarizing narratives were limited to only a few ecosystems, particularly “News & Hot Takes”. Misleading and selective reporting about firearm injury certainly begets more firearm injury (Beale, 2006; Garland, 2000; Jetter & Walker, 2018; Kaufman et al., 2020). Given the influence of the “New & Hot Takes” ecosystem, it’s likely that it drives the common misconception of firearm-related harms as random, unpreventable attacks by strangers (Anestis et al., 2021; Getman et al., 2018; Ye et al., 2022). This analysis emphasizes, however, that a focus on the extremes ignores the ways in which most Americans encounter, play with, and discuss real or imagined firearms (Downs, 2002).

Finally and most importantly, ample work on other public health topics - ranging from suicide prevention and bullying prevention (Uhls et al., 2021), to tobacco and e-cigarette use (Majmundar et al., 2021; Sargent et al., 2003), to vaccine uptake (Getman et al., 2018), to sexual harrassment (Huseth-Zosel et al., 2021), to firearm injury itself (Ojo et al., 2021) - demonstrates that cultural cognitions and behavior can be changed by combining data and narrative in the online world. In work on other public health problems, we see the importance of moving beyond the trope that one narrative is right, and another is wrong. Rather, awareness of the various narratives allows incorporation of facts - around actual patterns of injury and ways to protect from injury - into these online communities. With expanded qualitative and quantitative understanding of the narratives that specific sub-groups interact with, we can begin to develop interventions to shift norms, beliefs, and self-efficacy, in ways that are tailored to both ecosystem content and user group.

Limitations of this study include potential biases in the YouTube and internet search data, reflecting the short time-window, the single source of social media data, and the limited numbers of internet search providers and internet searches. Although the proprietary panel is supposed to reflect a diversity of American adults, it is possible that the user participation is biased in unknown ways. These unmeasured biases reinforce that this is a preliminary study, and should be reproduced with a larger number of social media sources and a wider array of internet searches. A further limitation is that although the study combined rigorous quantitative and qualitative methods, more nuanced inquiry into meaning and sentiment is needed; for example, advanced machine learning sentiment analysis could be conducted to better quantify the types and meaning of internet searches within the various ecosystems. Finally, and most importantly, this study was purely observational, with no measurement of actual behaviors or beliefs, no claim of causality between exposure and outcomes, and no intervention delivered.

## 5. Conclusion

In this study, we combined content analysis of online social networks with the search patterns of a representative sample of users to not only identify distinct narratives about firearms, but to also understand which segments of the population engaged with these narratives. Our results highlight the degree to which firearms are present in, and influence, Americans’ public and private life, in areas ranging from hobbies to entertainment to news and social events. Our results also highlight the diversity of people and experiences that are part of this multiplicity of narratives. As Americans experience firearms on YouTube, online searches and associated results, and then circle back again, a complex understanding of “firearms” and “firearm injury” is created. Future work should further define the types of narrative and information to which people are exposed, the demographic differences in narrative exposure, and how narratives of harm and safety can be incorporated into these pervasive online narrative ecosystems.

## Data Availability

All data are proprietary, anonymized databases of YouTube watching data.

## Acknowledgements/funding

We acknowledge Hayley Benson, Tayler Bunge and Vipassana Vijayarangan for their assistance with elements of study management and manuscript preparation.

Megan Ranney received funding in part from CDC R01 CE003267, NIH P20 GM139664, and NIH R24 HD087149 for this work. She is a Senior Strategic Advisor (volunteer) for AFFIRM at the Aspen Institute, and serves on the Board of Directors for the Nonviolence Institute in Providence, RI. Frederica R. Conrey is employed by Harmony Labs and Rory Smith was employed by Harmony Labs when this research was conducted.

