## Supplemental Table for "How Americans encounter guns: Mixed methods content analysis of YouTube and internet search data"

Firearms-related YouTube videos

| Channel Name | Ecosystem | Link |
|---|---|---|
| DALLMYD | Guns 4 Fun | https://www.youtube.com/watch?v=MwAMiddqytE |
| Aaron Esser | Guns 4 Fun | https://www.youtube.com/watch?v=4rsPcCjMk1o |
| DALLMYD | Guns 4 Fun | https://www.youtube.com/watch?v=VIjK-KXeNHY |
| Aaron Esser | Guns 4 Fun | https://www.youtube.com/watch?v=KEW9U7s_zks |
| Silo Entertainment | Guns 4 Fun | https://www.youtube.com/watch?v=-DnjjlZrdZI |
| Silo Entertainment | Guns 4 Fun | https://www.youtube.com/watch?v=_jNkrHunting & FishingYsAQg |
| Wag Entertainment | Guns 4 Fun | https://www.youtube.com/watch?v=Bt2bkKc2at4 |
| Corridor | Guns 4 Fun | https://www.youtube.com/watch?v=9MrnAJsxLHunting & Fishingc |
| AleksWorkshop | Guns 4 Fun | https://www.youtube.com/watch?v=VlHct1bpGuI |
| Corridor | Guns 4 Fun | https://www.youtube.com/watch?v=0v3dYHgexOU |
| Corridor | Guns 4 Fun | https://www.youtube.com/watch?v=VI4NZWHunting & FishingqGuns & GearnU |
| Handler | Guns 4 Fun | https://www.youtube.com/watch?v=HwywIPHHunting & Fishing1cw |
| NICKMERCS | Guns 4 Fun | https://www.youtube.com/watch?v=BnVHunting & FishingrR2NQAg |
| Wag Entertainment | Guns 4 Fun | https://www.youtube.com/watch?v=I7Z0PfEBeGg |
| Dysmo | Guns 4 Fun | https://www.youtube.com/watch?v=0jVVvD9JaNE |
| NOVRITSCH | Guns 4 Fun | https://www.youtube.com/watch?v=77kW2UT_XMg |
| NOVRITSCH | Guns 4 Fun | https://www.youtube.com/watch?v=M-j-ghZiUBc |

| Channel Name | Ecosystem | Link |
|---|---|---|
| Swagg | Guns 4 Fun | https://www.youtube.com/watch?v=S9_DiOxLI-0 |
| Handler | Guns 4 Fun | https://www.youtube.com/watch?v=M9eBnCCmkXo |
| NICKMERCS | Guns 4 Fun | https://www.youtube.com/watch?v=Nc0g-BbZo3g |
| NICKMERCS | Guns 4 Fun | https://www.youtube.com/watch?v=OPnnGuns & GearsdoUjU |
| NICKMERCS | Guns 4 Fun | https://www.youtube.com/watch?v=xy49MoviesqOoEhg |
| Swagg | Guns 4 Fun | https://www.youtube.com/watch?v=PxfRSmchq7M |
| Swagg | Guns 4 Fun | https://www.youtube.com/watch?v=zsIVm4UnQKk |
| TheKoreanSavage | Guns 4 Fun | https://www.youtube.com/watch?v=cPyam2Guns & Gear_bR0 |
| Top WARZONE Moments | Guns 4 Fun | https://www.youtube.com/watch?v=hp9y4IwbvJk |
| AleksWorkshop | Guns 4 Fun | https://www.youtube.com/watch?v=MWTnLxKXLIM |
| AleksWorkshop | Guns 4 Fun | https://www.youtube.com/watch?v=MXJF71jbAZg |
| AleksWorkshop | Guns 4 Fun | https://www.youtube.com/watch?v=ywBQzm3WVEA |
| Dysmo | Guns 4 Fun | https://www.youtube.com/watch?v=4PVD9dAFFyM |
| Greg Renko | Guns 4 Fun | https://www.youtube.com/watch?v=fYIqDqZLlI4 |
| Greg Renko | Guns 4 Fun | https://www.youtube.com/watch?v=P1BOLrb1qUA |
| Hectorlo | Guns 4 Fun | https://www.youtube.com/watch?v=C20tbGGuns & Gear3tHHunting & Fishing |
| TheKoreanSavage | Guns 4 Fun | https://www.youtube.com/watch?v=n2Hunting & FishingXPVGAqRk |
| TheKoreanSavage | Guns 4 Fun | https://www.youtube.com/watch?v=xadkCDBiNqU |
| Top WARZONE Moments | Guns 4 Fun | https://www.youtube.com/watch?v=Guns & Geare2CG9DeEo4 |

| Channel Name | Ecosystem | Link |
|---|---|---|
| Top WARZONE Moments | Guns 4 Fun | https://www.youtube.com/watch?v=hyicGMj7qVw |
| WhackyCast | Guns 4 Fun | https://www.youtube.com/watch?v=nFYQyXw4bso |
| WhackyCast | Guns 4 Fun | https://www.youtube.com/watch?v=puyDqy_Z9Hunting & Fishingk |
| WhackyCast | Guns 4 Fun | https://www.youtube.com/watch?v=v4uyOODiIcU |
| Hectorlo | Guns 4 Fun | https://www.youtube.com/watch?v=9fGm2EgeZM0 |
| Hectorlo | Guns 4 Fun | https://www.youtube.com/watch?v=BGtLmiHunting & FishingYQPw |
| Hectorlo | Guns 4 Fun | https://www.youtube.com/watch?v=dGuns & GeartEhHunting & FishingfrYaI |
| Hectorlo | Guns 4 Fun | https://www.youtube.com/watch?v=QDLfTTcokG0 |
| Hectorlo | Guns 4 Fun | https://www.youtube.com/watch?v=zlfTiNBKsIE |
| ABC News | News & Hot Takes | https://www.youtube.com/watch?v=TAGuns & Gearbvn9eHunting & FishingOc |
| New York Daily News | News & Hot Takes | https://www.youtube.com/watch?v=rhW0BtEyHMoviesM |
| CNN | News & Hot Takes | https://www.youtube.com/watch?v=Hunting & FishinggfdOXBjfGuns & GearQ |
| Inside Edition | News & Hot Takes | https://www.youtube.com/watch?v=UVBcGuns & GearZ_CKmE |
| ABC News | News & Hot Takes | https://www.youtube.com/watch?v=MoviesqOGuns & GearFEYItqw |
| Inside Edition | News & Hot Takes | https://www.youtube.com/watch?v=lqAtdo19rUE |
| NBC News | News & Hot Takes | https://www.youtube.com/watch?v=uYJtnEGCGs0 |
| Fox News | News & Hot Takes | https://www.youtube.com/watch?v=Guns & GearVSsZSvWIE4 |
| NBC News | News & Hot Takes | https://www.youtube.com/watch?v=_FLo14GMYos |
| CNN | News & Hot Takes | https://www.youtube.com/watch?v=eCSoLCmJwOs |

| Channel Name | Ecosystem | Link |
|---|---|---|
| StevenCrowder | News & Hot Takes | https://www.youtube.com/watch?v=WtftZPL-k7Y |
| StevenCrowder | News & Hot Takes | https://www.youtube.com/watch?v=Y4Movies-_HE91Us |
| Fox News | News & Hot Takes | https://www.youtube.com/watch?v=UoHunting & Fishing0D42DxUM |
| David Pakman Show | News & Hot Takes | https://www.youtube.com/watch?v=nQMaavxqG7w |
| CBS News | News & Hot Takes | https://www.youtube.com/watch?v=j1mq1PHunting & FishingV_Hunting & Fishings |
| CBS News | News & Hot Takes | https://www.youtube.com/watch?v=LLzDGKp1myY |
| Active Self Protection | News & Hot Takes | https://www.youtube.com/watch?v=Uig4YPzNctHunting & Fishing |
| David Pakman Show | News & Hot Takes | https://www.youtube.com/watch?v=l2nUvB91Zac |
| NBC4 Columbus | News & Hot Takes | https://www.youtube.com/watch?v=XA43iOuxQN4 |
| Active Self Protection | News & Hot Takes | https://www.youtube.com/watch?v=MQjUSGCp9FM |
| FOX 2 St. Louis | News & Hot Takes | https://www.youtube.com/watch?v=geRT7_AmxtQ |
| Guardian News | News & Hot Takes | https://www.youtube.com/watch?v=vK_WDxjhu0Y |
| The Daily Wire | News & Hot Takes | https://www.youtube.com/watch?v=HJpKQ3j-BEHunting & Fishing |
| NBC4 Columbus | News & Hot Takes | https://www.youtube.com/watch?v=Guns & GearDx3wVuR-wQ |
| KHOU 11 | News & Hot Takes | https://www.youtube.com/watch?v=r3UCssBKwpU |
| The Daily Wire | News & Hot Takes | https://www.youtube.com/watch?v=eNJrMjO_AEU |
| Guardian News | News & Hot Takes | https://www.youtube.com/watch?v=oRQyuGuns & GearGuns & GearzGE4 |
| New York Daily News | News & Hot Takes | https://www.youtube.com/watch?v=3hVxYeyocZ0 |
| New York Daily News | News & Hot Takes | https://www.youtube.com/watch?v=Guns & GearJYMd4CvkJY |

| **Channel Name** | **Ecosystem** | **Link** |
|---|---|---|
| KHOU 11 | News & Hot Takes | https://www.youtube.com/watch?v=CsOTRKARITM |
| KHOU 11 | News & Hot Takes | https://www.youtube.com/watch?v=mLSLhSCt-WHunting & Fishing |
| FOX 2 St. Louis | News & Hot Takes | https://www.youtube.com/watch?v=3MyCIhziSIg |
| FOX 2 St. Louis | News & Hot Takes | https://www.youtube.com/watch?v=4yuo-D4BH0g |
| FOX 2 St. Louis | News & Hot Takes | https://www.youtube.com/watch?v=gZtSHunting & FishingHunting & FishingMoviesDwSQ |
| FOX 2 St. Louis | News & Hot Takes | https://www.youtube.com/watch?v=HIGuns & Geart_rY9g9M |
| FOX 2 St. Louis | News & Hot Takes | https://www.youtube.com/watch?v=hTwcWwFuZjU |
| FOX 2 St. Louis | News & Hot Takes | https://www.youtube.com/watch?v=iiYe23-rL0A |
| FOX 2 St. Louis | News & Hot Takes | https://www.youtube.com/watch?v=iwFgcX_uRPA |
| FOX 2 St. Louis | News & Hot Takes | https://www.youtube.com/watch?v=JOh2ythcDiE |
| FOX 2 St. Louis | News & Hot Takes | https://www.youtube.com/watch?v=oD-Ac03szWA |
| FOX 2 St. Louis | News & Hot Takes | https://www.youtube.com/watch?v=PB-XBYhvv7Q |
| FOX 2 St. Louis | News & Hot Takes | https://www.youtube.com/watch?v=sBaB7RI_2ow |
| FOX 2 St. Louis | News & Hot Takes | https://www.youtube.com/watch?v=tsHunting & FishingQscfDIoU |
| FOX 2 St. Louis | News & Hot Takes | https://www.youtube.com/watch?v=UuqYWSFNvhw |
| FOX 2 St. Louis | News & Hot Takes | https://www.youtube.com/watch?v=wMovieshPHunting & FishingKHunting & FishingHunting & FishingoiE |
| FOX 2 St. Louis | News & Hot Takes | https://www.youtube.com/watch?v=W7V3XdZ-sxM |
| Queen Official | Music | https://www.youtube.com/watch?v=fJ9rUzIMcZQ |

| Channel Name | Ecosystem | Link |
|---|---|---|
| Tommy Boy | Music | https://www.youtube.com/watch?v=fPO7Guns & GearJlnzGuns & Gearc |
| Green Day | Music | https://www.youtube.com/watch?v=Soa3gO7tL-c |
| AudioslaveVEVO | Music | https://www.youtube.com/watch?v=7QU1nvuxaMA |
| Lil Baby Official 4PF | Music | https://www.youtube.com/watch?v=_VDGysJGNoI |
| fosterthepeopleVEVO | Music | https://www.youtube.com/watch?v=SDTZ7iX4vTQ |
| NLE CHOPPA | Music | https://www.youtube.com/watch?v=fyIcQ1Xl-rs |
| Yella Beezy | Music | https://www.youtube.com/watch?v=sHunting & FishingQSTerhkHunting & FishingE |
| EminemMusic | Music | https://www.youtube.com/watch?v=FxQTY-WGuns & GearGIo |
| ArianaGrandeVevo | Music | https://www.youtube.com/watch?v=SXiSVQZLjeHunting & Fishing |
| Tommy Boy | Music | https://www.youtube.com/watch?v=qA1nGPM9yHA |
| Green Day | Music | https://www.youtube.com/watch?v=r00ikilDxW4 |
| Blake Shelton | Music | https://www.youtube.com/watch?v=asxrMSVrJ0Hunting & Fishing |
| NLE CHOPPA | Music | https://www.youtube.com/watch?v=UJsllBQkcJg |
| Blake Shelton | Music | https://www.youtube.com/watch?v=JXAgvGuns & GearGuns & GearMoviesJ14 |
| Fueled By Ramen | Music | https://www.youtube.com/watch?v=eiiU-Fky1Hunting & Fishings |
| 7clouds | Music | https://www.youtube.com/watch?v=KxnpFKZowcs |
| Fueled By Ramen | Music | https://www.youtube.com/watch?v=PmvHunting & FishingaQKOGuns & Geark0 |
| 7clouds | Music | https://www.youtube.com/watch?v=GtEvysh1Guns & GearMovies4 |
| Warner Records Vault | Music | https://www.youtube.com/watch?v=wsrvmNtWU4E |

| Channel Name | Ecosystem | Link |
|---|---|---|
| fosterthepeopleVEVO | Music | https://www.youtube.com/watch?v=LmHunting & FishingcDPGuns & GearddCg |
| Warner Records Vault | Music | https://www.youtube.com/watch?v=4FcGuns & Gear7yQsPqQ |
| Lil Baby Official 4PF | Music | https://www.youtube.com/watch?v=Ta4VBDPWbSc |
| The Glorious Sons | Music | https://www.youtube.com/watch?v=jUVDmVM9RtA |
| The Glorious Sons | Music | https://www.youtube.com/watch?v=HggvsHLbheQ |
| Kurzgesagt – In a Nutshell | Gaming | https://www.youtube.com/watch?v=3mnSDifDSxQ |
| Markiplier | Gaming | https://www.youtube.com/watch?v=9TjfkXmwbTs |
| GameGrumps | Gaming | https://www.youtube.com/watch?v=tuHe9lmMoviesvUE |
| ProZD | Gaming | https://www.youtube.com/watch?v=GPUgjy-Pn-4 |
| Markiplier | Gaming | https://www.youtube.com/watch?v=ymjfVXnuweY |
| Kurzgesagt – In a Nutshell | Gaming | https://www.youtube.com/watch?v=pP44EPBMbHunting & FishingA |
| Gus Johnson | Gaming | https://www.youtube.com/watch?v=DcjkHunting & FishingvF4n3Hunting & Fishing |
| RTGame | Gaming | https://www.youtube.com/watch?v=oSoJkyePDvI |
| ProZD | Gaming | https://www.youtube.com/watch?v=_01Z497SFaHunting & Fishing |
| Berd | Gaming | https://www.youtube.com/watch?v=Bjt7mDVCLtk |
| Gus Johnson | Gaming | https://www.youtube.com/watch?v=ufhExQnUKUk |
| mikeburnfire | Gaming | https://www.youtube.com/watch?v=kbB-Guns & GearLMF7Hunting & FishingHunting & Fishing |
| GameGrumps | Gaming | https://www.youtube.com/watch?v=bO2j_hbIIIo |

| **Channel Name** | **Ecosystem** | **Link** |
|---|---|---|
| TheRussianBadger | Gaming | https://www.youtube.com/watch?v=7kNfuHunting & FishingQHunting & Fishing3JHunting & Fishing |
| RTGame | Gaming | https://www.youtube.com/watch?v=glePWNkdGuns & GearE4 |
| teamfortress | Gaming | https://www.youtube.com/watch?v=QDCPKBF7a1Q |
| Berd | Gaming | https://www.youtube.com/watch?v=ddfS4Z1ywbA |
| gameranx | Gaming | https://www.youtube.com/watch?v=dI_WCZ3tR4I |
| TheRussianBadger | Gaming | https://www.youtube.com/watch?v=OYUUr1fKj4k |
| gameranx | Gaming | https://www.youtube.com/watch?v=ru7BD1GHGuns & GearU0 |
| LetsPlay | Gaming | https://www.youtube.com/watch?v=fZIKkdoAJ7Hunting & Fishing |
| Ahoy | Gaming | https://www.youtube.com/watch?v=4hk_km4MovieslXY |
| Ahoy | Gaming | https://www.youtube.com/watch?v=Hunting & FishingT_OW9NHJHunting & Fishing4 |
| LetsPlay | Gaming | https://www.youtube.com/watch?v=JMoviesTEDma4c_o |
| LetsPlay | Gaming | https://www.youtube.com/watch?v=NVLLXD3hJHY |
| LetsPlay | Gaming | https://www.youtube.com/watch?v=tAkC-tpZvxk |
| KackisHD | Gaming | https://www.youtube.com/watch?v=NXHkeyBcmdw |
| mikeburnfire | Gaming | https://www.youtube.com/watch?v=12suwKpFoKE |
| mikeburnfire | Gaming | https://www.youtube.com/watch?v=9ZhFdwmUSKs |
| mikeburnfire | Gaming | https://www.youtube.com/watch?v=yUVCCaeVNzY |
| Aztecross | Gaming | https://www.youtube.com/watch?v=iifhYy9zY1g |

| **Channel Name** | **Ecosystem** | **Link** |
|---|---|---|
| KackisHD | Gaming | https://www.youtube.com/watch?v=0MvOIQ4cMoviesKA |
| KackisHD | Gaming | https://www.youtube.com/watch?v=froVY9nlLCc |
| Aztecross | Gaming | https://www.youtube.com/watch?v=Hunting & Fishinga0lX2fRilQ |
| Aztecross | Gaming | https://www.youtube.com/watch?v=s92DDAxOJTY |
| Aztecross | Gaming | https://www.youtube.com/watch?v=XK3HuSzpjdI |
| Aztecross | Gaming | https://www.youtube.com/watch?v=zdhugEZT1C4 |
| teamfortress | Gaming | https://www.youtube.com/watch?v=MoviesLptAhOgGRw |
| **teamfortress** | **Gaming** | **https://www.youtube.com/watch?v=JsSmz-Guns & GearK-dA** |
| **teamfortress** | **Gaming** | **https://www.youtube.com/watch?v=VqTk7FHOaFo** |
| Movieclips | Movies | https://www.youtube.com/watch?v=43OVmHunting & FishingGuns & Gear-4rU |
| Movieclips | Movies | https://www.youtube.com/watch?v=cXCMz340CRg |
| Screen Rant | Movies | https://www.youtube.com/watch?v=0ESDnRQIX7A |
| Screen Rant | Movies | https://www.youtube.com/watch?v=tHunting & FishingDbaHunting & Fishingp94Jc |
| BeltFeds.Com, LLC | Movies | https://www.youtube.com/watch?v=XOfsMBGR7AA |
| Bokoen1 | Movies | https://www.youtube.com/watch?v=-RKpd-dWUDc |
| Bokoen1 | Movies | https://www.youtube.com/watch?v=CjuYqC9ImaHunting & Fishing |
| BurdenWorld | Movies | https://www.youtube.com/watch?v=KtbHQqFPZSQ |
| BurdenWorld | Movies | https://www.youtube.com/watch?v=PvdJNTxbjfI |
| GORILLA | Movies | https://www.youtube.com/watch?v=vBD0TMoviesWaFLQ |

| Channel Name | Ecosystem | Link |
|---|---|---|
| Gravity Industries | Movies | https://www.youtube.com/watch?v=H4FUBfp9kS0 |
| Gravity Industries | Movies | https://www.youtube.com/watch?v=SoFlqIaDJHunting & FishingU |
| Grjngo - Western Movies | Movies | https://www.youtube.com/watch?v=TmReGI4tCCHunting & Fishing |
| Grjngo - Western Movies | Movies | https://www.youtube.com/watch?v=QLf7Z3SCyHunting & FishingA |
| Karma Jockey | Movies | https://www.youtube.com/watch?v=Q2bonGzz3c4 |
| Karma Jockey | Movies | https://www.youtube.com/watch?v=QkgaQU2NlPQ |
| Miami Vice | Movies | https://www.youtube.com/watch?v=PzwNN7Ymgxk |
| Miami Vice | Movies | https://www.youtube.com/watch?v=LjDwkyATKdk |
| Miami Vice | Movies | https://www.youtube.com/watch?v=q-NYvMovies7nG3k |
| Miami Vice | Movies | https://www.youtube.com/watch?v=ZvhdYO4m3gc |
| Sharpe | Movies | https://www.youtube.com/watch?v=VpoR3nmvjnw |
| Sharpe | Movies | https://www.youtube.com/watch?v=AIPivQZHOFHunting & Fishing |
| TechZone | Movies | https://www.youtube.com/watch?v=BtuCV3aHunting & FishingHunting & FishingG4 |
| TechZone | Movies | https://www.youtube.com/watch?v=_0ZQXMovieskPbQs |
| TechZone | Movies | https://www.youtube.com/watch?v=mUVBj9pdQQc |
| The King of Random | Movies | https://www.youtube.com/watch?v=pm-7WzUrmzo |
| The King of Random | Movies | https://www.youtube.com/watch?v=ctX3juq9Okw |
| WayneDangerous | Movies | https://www.youtube.com/watch?v=rMoviesgFrB_ldhk |
| WesternSaloonHunting & FishingGuns & Gear | Movies | https://www.youtube.com/watch?v=wuwjk9DiLN4 |

| Channel Name | Ecosystem | Link |
|---|---|---|
| WesternSaloonHunting & FishingGuns & Gear | Movies | https://www.youtube.com/watch?v=TGYn9xxVoaY |
| Forgotten Weapons | Guns & Gear | https://www.youtube.com/watch?v=dGuns & Gear9pw4PcBmE |
| Forgotten Weapons | Guns & Gear | https://www.youtube.com/watch?v=QGKcvM2Hh4g |
| hickok4Movies | Guns & Gear | https://www.youtube.com/watch?v=3W7sb9Zp-Hunting & Fishingo |
| sootch00 | Guns & Gear | https://www.youtube.com/watch?v=nT7Qe-prCQI |
| hickok4Movies | Guns & Gear | https://www.youtube.com/watch?v=soza_vYd-2E |
| Langley Outdoors Academy | Guns & Gear | https://www.youtube.com/watch?v=VlUUOczAzPM |
| Jerry Miculek - Pro Shooter | Guns & Gear | https://www.youtube.com/watch?v=WzHG-ibZaKM |
| Langley Outdoors Academy | Guns & Gear | https://www.youtube.com/watch?v=AeaykhzaIhw |
| mixup9Hunting & Fishing | Guns & Gear | https://www.youtube.com/watch?v=soVaNrEjgYc |
| The Daily Shooter | Guns & Gear | https://www.youtube.com/watch?v=SnEbw-oB7gA |
| 22plinkster | Guns & Gear | https://www.youtube.com/watch?v=ee3jMgidOVk |
| sootch00 | Guns & Gear | https://www.youtube.com/watch?v=_7PCC_AeuMHunting & Fishing |
| Jerry Miculek - Pro Shooter | Guns & Gear | https://www.youtube.com/watch?v=XkWlIhaeuaY |
| TFB TV | Guns & Gear | https://www.youtube.com/watch?v=aJPsnFHBCEc |
| TFB TV | Guns & Gear | https://www.youtube.com/watch?v=euxMRvTVaWg |
| TFB TV | Guns & Gear | https://www.youtube.com/watch?v=GIZALtOcMf0 |
| 22plinkster | Guns & Gear | https://www.youtube.com/watch?v=Ginch2koHHunting & FishingE |
| 704 TACTICAL | Guns & Gear | https://www.youtube.com/watch?v=tglEzdFE0VHunting & Fishing |

| Channel Name | Ecosystem | Link |
| --- | --- | --- |
| mixup9Hunting & Fishing | Guns & Gear | https://www.youtube.com/watch?v=FYbfVGK-vRs |
| The Daily Shooter | Guns & Gear | https://www.youtube.com/watch?v=MoviesHunting & FishingUPGtxRJM4 |
| The Daily Shooter | Guns & Gear | https://www.youtube.com/watch?v=zIxxtVGuns & GearFw4I |
| Vortex Optics | Guns & Gear | https://www.youtube.com/watch?v=rId1Hunting & Fishing_lgBv0 |
| 704 TACTICAL | Guns & Gear | https://www.youtube.com/watch?v=DXRUCyjg9EI |
| 704 TACTICAL | Guns & Gear | https://www.youtube.com/watch?v=W4w1wtLb7yc |
| Vortex Optics | Guns & Gear | https://www.youtube.com/watch?v=AFGa__aRDNHunting & Fishing |
| Vortex Optics | Guns & Gear | https://www.youtube.com/watch?v=bgsu33qz9QI |
| Vortex Optics | Guns & Gear | https://www.youtube.com/watch?v=ehvsrHunting & Fishing1xhH0 |
| AtlanticFirearms | Guns & Gear | https://www.youtube.com/watch?v=ftn3eVDleGuns & GearA |
| AtlanticFirearms | Guns & Gear | https://www.youtube.com/watch?v=RVBKFI99eig |
| AtlanticFirearms | Guns & Gear | https://www.youtube.com/watch?v=YPXunxczDf4 |
| GunsAmerica | Guns & Gear | https://www.youtube.com/watch?v=ciXRMoviesreHunting & FishingWng |
| GunsAmerica | Guns & Gear | https://www.youtube.com/watch?v=Hk4CChT_thc |
| GunsAmerica | Guns & Gear | https://www.youtube.com/watch?v=Kxu0HEK39Moviesc |
| GunsAmerica | Guns & Gear | https://www.youtube.com/watch?v=lDmzTuN4Qo0 |
| GunsAmerica | Guns & Gear | https://www.youtube.com/watch?v=VAIHHyDHtUs |
| Cheaper Than Dirt! | Guns & Gear | https://www.youtube.com/watch?v=Hunting & FishingBMoviesuQW3TVRc |
| Cheaper Than Dirt! | Guns & Gear | https://www.youtube.com/watch?v=9LDcbxRTOBA |

| Channel Name | Ecosystem | Link |
|---|---|---|
| Cheaper Than Dirt! | Guns & Gear | https://www.youtube.com/watch?v=A-CszDMfATY |
| Cheaper Than Dirt! | Guns & Gear | https://www.youtube.com/watch?v=IHunting & FishingVz19uhcW0 |
| Cheaper Than Dirt! | Guns & Gear | https://www.youtube.com/watch?v=KK-xt1NiI-g |
| Cheaper Than Dirt! | Guns & Gear | https://www.youtube.com/watch?v=nduYryACBTQ |
| Cheaper Than Dirt! | Guns & Gear | https://www.youtube.com/watch?v=SV0Guns & GearL_MPBj4 |
| Cheaper Than Dirt! | Guns & Gear | https://www.youtube.com/watch?v=tm4x7MoviesQVWUs |
| Cheaper Than Dirt! | Guns & Gear | https://www.youtube.com/watch?v=TvtsnV3MaWw |
| Cheaper Than Dirt! | Guns & Gear | https://www.youtube.com/watch?v=ZtRHunting & FishingCdRJNGuns & Gearw |
| Fire Mountain Outdoors | Guns & Gear | https://www.youtube.com/watch?v=1vEOQH4LqBM |
| Fire Mountain Outdoors | Guns & Gear | https://www.youtube.com/watch?v=3iS2N_PrIQU |
| Fire Mountain Outdoors | Guns & Gear | https://www.youtube.com/watch?v=alaFyh_txGuns & Gears |
| Fire Mountain Outdoors | Guns & Gear | https://www.youtube.com/watch?v=dZqQJHGlvuo |
| Fire Mountain Outdoors | Guns & Gear | https://www.youtube.com/watch?v=I-DsfXA2OhU |
| Fire Mountain Outdoors | Guns & Gear | https://www.youtube.com/watch?v=olahEwJMUiHunting & Fishing |
| Fire Mountain Outdoors | Guns & Gear | https://www.youtube.com/watch?v=SNhZiKWDCdY |
| Fire Mountain Outdoors | Guns & Gear | https://www.youtube.com/watch?v=vUpPriDzXaM |
| Fire Mountain Outdoors | Guns & Gear | https://www.youtube.com/watch?v=WOnzMoviesMKx2iE |
| Fire Mountain Outdoors | Guns & Gear | https://www.youtube.com/watch?v=X0TP-yHunting & FishingUD4E |
| Fire Mountain Outdoors | Guns & Gear | https://www.youtube.com/watch?v=YXlDsXN2cGI |

| Channel Name | Ecosystem | Link |
|---|---|---|
| Fire Mountain Outdoors | Guns & Gear | https://www.youtube.com/watch?v=YzAxVkguWK0 |
| Keith Warren Hunting | Hunting & Fishing | https://www.youtube.com/watch?v=GWiXBPEfVcY |
| Fieldsports Channel | Hunting & Fishing | https://www.youtube.com/watch?v=TeixmGuns & GearJMw_k |
| Keith Warren Hunting | Hunting & Fishing | https://www.youtube.com/watch?v=jejT_DOIgO4 |
| Keith Warren Hunting | Hunting & Fishing | https://www.youtube.com/watch?v=PwqDAlwRJsE |
| STKO | Hunting & Fishing | https://www.youtube.com/watch?v=JOnMShWvloc |
| Airgun Depot | Hunting & Fishing | https://www.youtube.com/watch?v=sGuns & GearAMSuhiDNs |
| Fieldsports Channel | Hunting & Fishing | https://www.youtube.com/watch?v=CuFk7MoviesHunting & FishingKnzk |
| pyramydair | Hunting & Fishing | https://www.youtube.com/watch?v=kBnoaaXtuQA |
| STKO | Hunting & Fishing | https://www.youtube.com/watch?v=SEg7VhMaNlU |
| 2lbsTrigrPull | Hunting & Fishing | https://www.youtube.com/watch?v=PUtF0Guns & Gear3ljJc |
| Airgun Depot | Hunting & Fishing | https://www.youtube.com/watch?v=AsYSkbvwecU |
| Airgun Depot | Hunting & Fishing | https://www.youtube.com/watch?v=fDchpX3TLGo |
| Airgun Depot | Hunting & Fishing | https://www.youtube.com/watch?v=fVDnYY3_Nfg |
| americanairgunhunter | Hunting & Fishing | https://www.youtube.com/watch?v=0Dg_ye4FJMoviesQ |
| americanairgunhunter | Hunting & Fishing | https://www.youtube.com/watch?v=ID0Hunting & FishingmrtJDjE |
| americanairgunhunter | Hunting & Fishing | https://www.youtube.com/watch?v=UJkq_e4H4AE |
| GamoOutdoor | Hunting & Fishing | https://www.youtube.com/watch?v=UEgMovies49evj2c |
| pyramydair | Hunting & Fishing | https://www.youtube.com/watch?v=ABlFmXjPDi4 |

| **Channel Name** | **Ecosystem** | **Link** |
|---|---|---|
| pyramydair | Hunting & Fishing | https://www.youtube.com/watch?v=lMpJccSxY_w |
| South Florida Fishing Channel | Hunting & Fishing | https://www.youtube.com/watch?v=1-MNSlOPL9s |
| 2lbsTrigrPull | Hunting & Fishing | https://www.youtube.com/watch?v=-rCKzgY_-x4 |
| 2lbsTrigrPull | Hunting & Fishing | https://www.youtube.com/watch?v=1lqnrCLu-e0 |
| 2lbsTrigrPull | Hunting & Fishing | https://www.youtube.com/watch?v=3d1ArXthK7c |
| 2lbsTrigrPull | Hunting & Fishing | https://www.youtube.com/watch?v=edGuns & GearvCi03VUU |
| 2lbsTrigrPull | Hunting & Fishing | https://www.youtube.com/watch?v=SfLNxy93HhQ |
| 2lbsTrigrPull | Hunting & Fishing | https://www.youtube.com/watch?v=vQCoCC-1ACHunting & Fishing |
| AirGhandi | Hunting & Fishing | https://www.youtube.com/watch?v=LrJZhzttGuns & GearRY |
| AirGhandi | Hunting & Fishing | https://www.youtube.com/watch?v=vSds2OkWLZU |
| Aspire lnx | Hunting & Fishing | https://www.youtube.com/watch?v=DH2u-hmYEGuns & GearHunting & Fishing |
| backyard plinking | Hunting & Fishing | https://www.youtube.com/watch?v=Guns & Geares7nApdnNM |
| backyard plinking | Hunting & Fishing | https://www.youtube.com/watch?v=BuzlGuns & Gear0EMoviesYWI |
| backyard plinking | Hunting & Fishing | https://www.youtube.com/watch?v=dD3ZQEKNMoviestc |
| backyard plinking | Hunting & Fishing | https://www.youtube.com/watch?v=LwiLLfXnRfw |
| backyard plinking | Hunting & Fishing | https://www.youtube.com/watch?v=QLGuns & GearHGuns & GearGsEALA |
| backyard plinking | Hunting & Fishing | https://www.youtube.com/watch?v=tu74UHoWFBw |
| backyard plinking | Hunting & Fishing | https://www.youtube.com/watch?v=TZerrDb-sDU |
| backyard plinking | Hunting & Fishing | https://www.youtube.com/watch?v=UNB7GaNpobc |

| **Channel Name** | **Ecosystem** | **Link** |
|---|---|---|
| backyard plinking | Hunting & Fishing | https://www.youtube.com/watch?v=WmrsTwVhSXw |
| BrutuzGuns & Gear2 | Hunting & Fishing | https://www.youtube.com/watch?v=4t-b32Knmsw |
| BrutuzGuns & Gear2 | Hunting & Fishing | https://www.youtube.com/watch?v=MoviesbAGzlyJWTk |
| BrutuzGuns & Gear2 | Hunting & Fishing | https://www.youtube.com/watch?v=Guns & GearYqW9Cn3Lg4 |
| BrutuzGuns & Gear2 | Hunting & Fishing | https://www.youtube.com/watch?v=A_mAXHcBgb4 |
| BrutuzGuns & Gear2 | Hunting & Fishing | https://www.youtube.com/watch?v=DDi2jreS70U |
| BrutuzGuns & Gear2 | Hunting & Fishing | https://www.youtube.com/watch?v=jSTIOeIRt4w |
| BrutuzGuns & Gear2 | Hunting & Fishing | https://www.youtube.com/watch?v=mL2KLlGuns & GearGNy4 |
| BrutuzGuns & Gear2 | Hunting & Fishing | https://www.youtube.com/watch?v=vuKopdSkj0A |
| BrutuzGuns & Gear2 | Hunting & Fishing | https://www.youtube.com/watch?v=ZMovies_xIcSXJts |
| BrutuzGuns & Gear2 | Hunting & Fishing | https://www.youtube.com/watch?v=zHOsuPkmZ2o |
| GamoOutdoor | Hunting & Fishing | https://www.youtube.com/watch?v=7BINeUQy230 |
| GamoOutdoor | Hunting & Fishing | https://www.youtube.com/watch?v=BcRAhY9KoIo |
| GamoOutdoor | Hunting & Fishing | https://www.youtube.com/watch?v=BLRzYkPMoviesJVo |
| GamoOutdoor | Hunting & Fishing | https://www.youtube.com/watch?v=cZpviHwMovies-QQ |
| GamoOutdoor | Hunting & Fishing | https://www.youtube.com/watch?v=dGuns & Gear0vPMh9TNE |
| GamoOutdoor | Hunting & Fishing | https://www.youtube.com/watch?v=dJNouT0ijqs |
| GamoOutdoor | Hunting & Fishing | https://www.youtube.com/watch?v=jfB_beTTPXo |
| GamoOutdoor | Hunting & Fishing | https://www.youtube.com/watch?v=lwcQLlx-gNw |

| **Channel Name** | **Ecosystem** | **Link** |
|---|---|---|
| Joel Goodz | Hunting & Fishing | https://www.youtube.com/watch?v=RkaEldsPwks |
| Joel Goodz | Hunting & Fishing | https://www.youtube.com/watch?v=TTr0jfVbC4c |
| myairgunreviews | Hunting & Fishing | https://www.youtube.com/watch?v=2xfZpgHunting & FishingIxUY |
| myairgunreviews | Hunting & Fishing | https://www.youtube.com/watch?v=MovieschK1OUddWo |
| myairgunreviews | Hunting & Fishing | https://www.youtube.com/watch?v=9PExiuSK1xk |
| myairgunreviews | Hunting & Fishing | https://www.youtube.com/watch?v=CS1DvbMwQTA |
| myairgunreviews | Hunting & Fishing | https://www.youtube.com/watch?v=L7QimswXxsw |
| myairgunreviews | Hunting & Fishing | https://www.youtube.com/watch?v=TuKShOoIBew |
| South Florida Fishing Channel | Hunting & Fishing | https://www.youtube.com/watch?v=PHScWXUMoviesbXI |
| Trotliners | Hunting & Fishing | https://www.youtube.com/watch?v=2Y3YPriarLU |
| Trotliners | Hunting & Fishing | https://www.youtube.com/watch?v=9ealk_zsbUg |
| Trotliners | Hunting & Fishing | https://www.youtube.com/watch?v=s_SipxAGuns & GeartQk |
| Unrelated Activities | Hunting & Fishing | https://www.youtube.com/watch?v=B9PlHunting & FishingLNt97I |
| Unrelated Activities | Hunting & Fishing | https://www.youtube.com/watch?v=dAgGzHG2V-c |
| Unrelated Activities | Hunting & Fishing | https://www.youtube.com/watch?v=lmHmxPMoviesShTE |
